# On Dynamical Analysis of the Data-Driven SIR model (COVID-19 Outbreak in Indonesia)

**DOI:** 10.1101/2020.06.22.20137810

**Authors:** Albert Sulaiman

## Abstract

An archipelago country such as Indonesia has a different beginning of the outbreak, therefore the management of epidemics not uniform. For this reason, the results in the data of confirmed cases COVID-19 to fluctuate and difficult to predict. We use the data-driven SIR model to analyze the dynamics and behavior of the evolution of the disease. We run the data-driven SIR model gradually and found that there are shifting of the peak and the distance of saturation point. We found that a transmission acceleration of the outbreak occurring in Indonesia where it could be seen from increasing of the time the saturation and the confirmed cases. It is finally argued that a new parameter can be used to guidance the condition when the new normal begins.

## 1 Introduction

Nowadays, the coronavirus (Covid-19) continues to widen and gives a large social disruption around the world. Until the virus vaccine discovered, the epidemic ended when humans as a host’s contacts are immune (herd immunity). Although there is no vaccine, the viruses spreading can be prevented by wear face masks, social distancing and stay at home. This means that the evolution of diseases can be predicted [1]. A lot of researchers have done a prediction of the spreading and the end of the epidemic of Covid-19 applied around the world to a particular country [2, 3, 4, 5, 6, 7, 8, 9]. The model predictions essentially separate into two parts. First, the model based on data where this approach is known as a statistical model. This approach with various algorithms has successfully applied in some countries [3, 4, 9, 10, 11, 12]. The second approach based on the deterministic epidemic model known as susceptible-infectious-recovery (SIR) with its variation [13, 14, 15, 16].

Work with a deterministic model has an advantage i.e we can make certain scenarios as quarantine, the lockdown area, and others. This scenario can be used to manage the disease. For instance, by using the *SEIQ*_*n*_*R* model Misra et al proposed two kinds of quarantine (*Q*_*n*_) models i.e home isolation and hospital quarantine. They show that home isolation and quarantine to hospitals are the recommended way in preventing the spreading of this diseases [17]. The SEIQR model was applied to several cities simultaneously (n-compartment model) has done where effect quarantine reduces the infection significantly [19]. Another variance of the SEIR model is taking into account the death kinetics law which describes the patient either recovers or dies after some characteristic time [18]. This approach applies very well to a particular state but not for other countries.

A lot of model predictions have been developed, but essentially the model used is the SIR model. Although simple, the SIR model is most appropriate for predictions based on the data rather than the more complex models [20]. A model using the data as the main factors to predict something called the data-driven model. With a simple and data driven model based on the physics of natural growth algorithm, the time-dependent transmission rate can be determined and used for tracking coronavirus outbreak [21]. The data-driven model also useful for short term (real-time) forecasts and risk assessment of an ongoing outbreak [22].

Indonesia is an archipelago that had intro outbreak is not the same so that the management of epidemics like partial lockdown, quarantine is not uniform. This resulted in the data of confirmed cases COVID-19 be also fluctuated and difficult to predict. Some research institutes in Indonesia predicted at the beginning of the outbreak (3-2-2020) were generally the peak disease will occur in May [25]. But in reality, until early June confirmed cases still happen and tended to increase from day today. Required an adaptive model that is very important to decision-making. Paper discussed an analysis of the dynamics of the development of confirmed cases in Indonesia based on an adaptive called data-driven model. The detail derivation of the model is given in sec-2 and an analysis of cases in Indonesia represented in the following chapter. The conclusion gives a highlight of the results and suggestions for reducing the outbreak of COVID-19 disease.

## 2 The Data-Driven SIR model and Batista Code

The most common epidemic model is called SIR were consisting of three parameters *S, I*, and *R* for (Susceptible, Infectious, and Recovery) which are representative of a susceptible person contracting, people are infected and the person who recovering respectively [26]. The order of the letters shows the process, so *S* − *I* − *R* stated the process of vulnerable people and infected and finally healed. So far, a SIR model is used to an infectious disease is transmitted from human to human, where the recovery of giving resistance has a long time the sustainability such as measles, smallpox, and rubella. Recovery here can be defined as healed, immune, or die.

This model assumes that the (*N*) located within the enclosed system (*S* + *I* + *R* = *N*) where the population excluding migration and into the city. This condition advantageous because we are dealing with two parameters of *S, I*, and *R*. A second assumption is the average infected people (infection rate) proportional with a number of a susceptible person and the average frequency of who is recovering to proportional with infected people. By using this assumption the SIR model is expressed in the form of differential equations as follows. [26, 27],

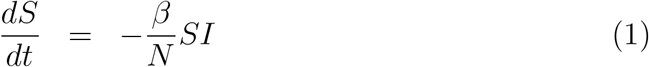

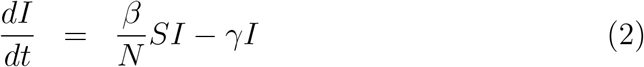

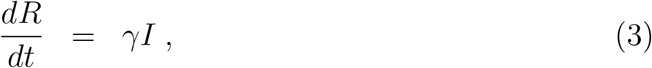

where *β* is the average number of the contact person^−1^ day^−1^, *γ* is individual infected rate. The second assumption, quantitatively shows that the dynamics of humans infected hanging from the ratio between *β* and *γ* namely *r* = *β/γ* known as basic reproduction number or shortened reproduction number. This can be derived from the second time the probability of becoming infected or often called the second wave infection. For example, influenza viruses even though they were recovery but can be infected, so the SIR model converted into the SIS model.From both the parameters define contact time *t*_*c*_ = *β*^−1^ and time of individual infected (incubation periods) or known as the average infectious period *t*_*r*_ = *γ*^−1^. From the simple model develop into the complex model for closer to the fields.

As mentioned above that the SIR model is built by a nonlinear differential equation with two unknown coefficients. In running the models hence these coefficients should be determined. If the condition of the outbreak is finished then a method of the curve fitting can be used to determine the coefficients of *β* and *γ*. If the coefficient of known them the evolution of epidemic can be seen in a proper possible way including when pandemic ended. How is if the condition of being held? How a decisive manner of the coefficients above? Batista develops the method and Matlab code to answer this question [28] that we called the Batista’s code. The method begins with fitting of the data by the equation represented the end of the outbreak. If the initial conditions expressed by (*S*_0_, *I*_0_, *R*_0_) substituting the third equation into the first of Eq-3 yields,

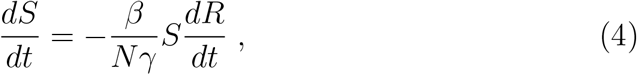

or we wrote to be, 

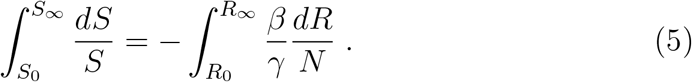

The integration of this equation yields,

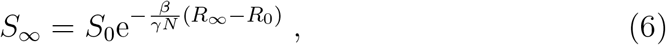

A sign ∞ is a condition where the epidemic has been ended. In the final of the outbreak (*I* = 0) there is no longer any infected so that (*N* = *S*_∞_ + *R*_∞_. By using Eq-6 then we got final value of recovery (*R*_∞_) namely,

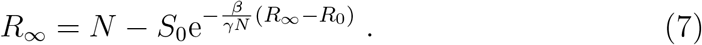

This equation used to estimate the coefficients of *β* and *γ* with the initial conditions *S*_0_, *I*_0_ and *R*_0_ from the field data.

## 3 Result and Discussion

The first confirmed case of coronavirus disease 2019 in Indonesia on 2 March 2020 and by 9 April the pandemic had spread to 34 provinces and still increase of new cases. The number of confirmed cases (*C* = *I* +*R*) and confirmed rate using data updated up to June 9 (https://www.kompas.com/covid-19/),(https://covid19.go.id/) are depicted in Fig-1. The highest confirmed cases that occur in Jakarta (the capital city of Indonesia) is about 23.68% of total cases followed by East Java province 20.3%, South Sulawesi 7.23%, West Java 6.91 % and Central Java 5.21%. Interestingly, the number of cases is not linear dependent on the distance from Jakarta where the first case began. Based on the distance, closest to Jakarta is West Java, Central Java, and East Java. The south Sulawesi province located in difference island with Jakarta and have the largest population in the eastern part of Indonesia. Started on 23 April 2020, Jakarta has approved large-scale social restrictions and followed by another province. From Fig.1 it seems on may 21 cases per day looked to start to be at the peak of the epidemic so that in early June some provinces have started to loosen policy large-scale social restrictions.

**Figure 1:**
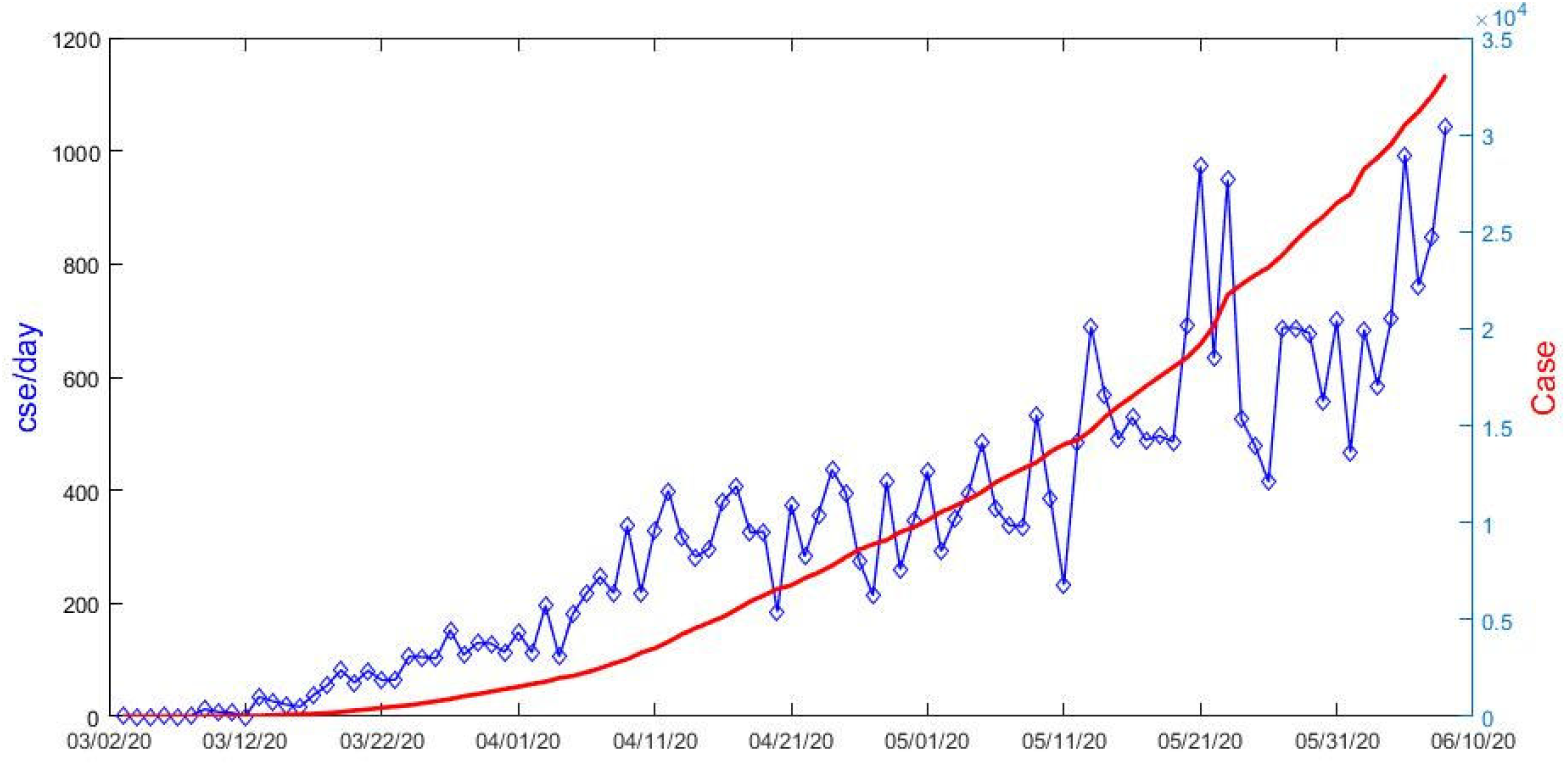
The number of confirmed cases (*C* = *I* + *R*) and confirmed rate using data updated from March 2 up to June 9. (https://www.kompas.com/covid-19/),(https://covid19.go.id/)

For analysis the peak of diseases, we run the data-driven SIR model which has to define it in Sec-2 gradually. The results of the calculation are shown in Fig.2. The figure shows that a comparison of the model and all data sets (confirmed cases) have high *R*^2^ *>* 0.98. The calculation of the model from the beginning of the outbreak until May 5 is expressed in Fig-2a. The simulation estimates that the outbreak will end on the date of June 9 where the top of outbreak happened to on April 20 (see figure the on the right hand). Fig-2b simulation until May 19 estimates that the outbreak will end on the date of June 29 where the top of the outbreak happened to on May 10. The simulation until May 28 shows the outbreak end on the date of September 17 where the peak of the outbreak occurs on May 21 see Fig-2c. Finally, the simulation until June 9 predict the saturation point occurs longer than September 17 and the peak of the epidemic occurs on June 6.

**Figure 2:**
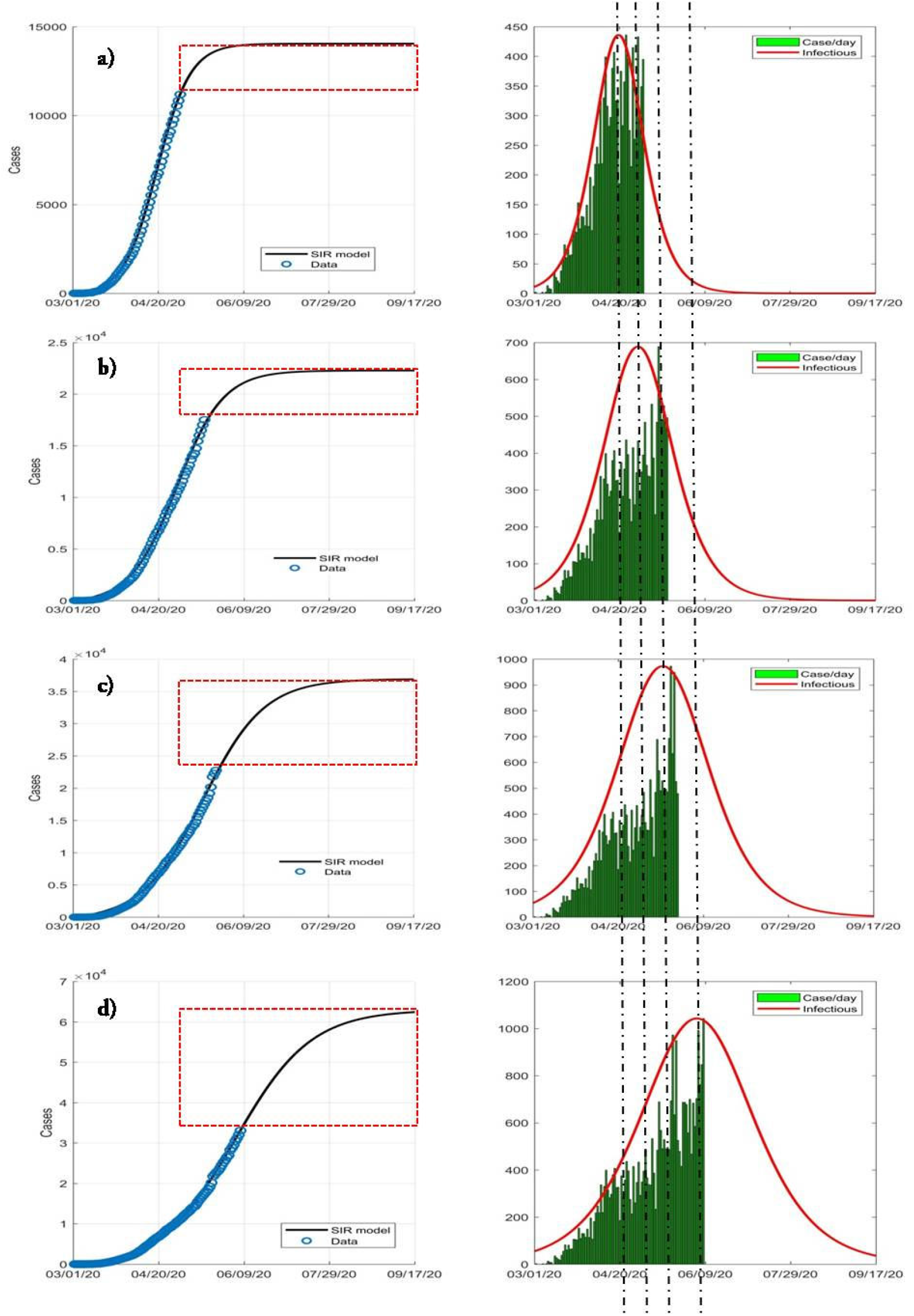
Simulation of the data-driven SIR gradually. a) The calculation of the model from the beginning of the outbreak until May 5, b) calculation until May 19, c) calculation until May 28 and d) calculation until June 9. On the left side, the black line is the confirmed case data and the blue circle is model prediction. The right side describes a comparison between cases per day and the Infectious model. The red box describes the difference between saturation time and the number of cases. The line-dot gives the shifting of the Infectious peak.

We see that there are clearly shifting of the peak and the distance of saturation point. This shows that the outbreak prediction at the current time is hard to do. In several countries with discipline and perspicuity regulation, this method is quite appropriate [7, 15]. They shows that implementation of stronger interventions to society could accelerate the complete recovery Heilongjiang province, China. Elsewhere, the data-driven simplified SIR model adequately reproduces outbreak dynamics qualitatively and quantitatively so that it can be used for predictive estimations [23, 24]. In Indonesia with many islands and early outbreak not uniforms, prediction overall difficult Fig-3. But in general, from Fig-2, there is a linear relationship between the time the saturation and the confirmed cases which are expressed in Fig-4. This linear regression had a good correlation coefficient is about 0.79 with gradient 250 of confirmed cases per day saturation. The interpretation is the condition of being concerned because there is an acceleration in the transmission of the outbreak in Indonesia. This condition is not out of these policies to loosen large-scale social restrictions in early June so that there an increasing number of cases every day.

**Figure 3:**
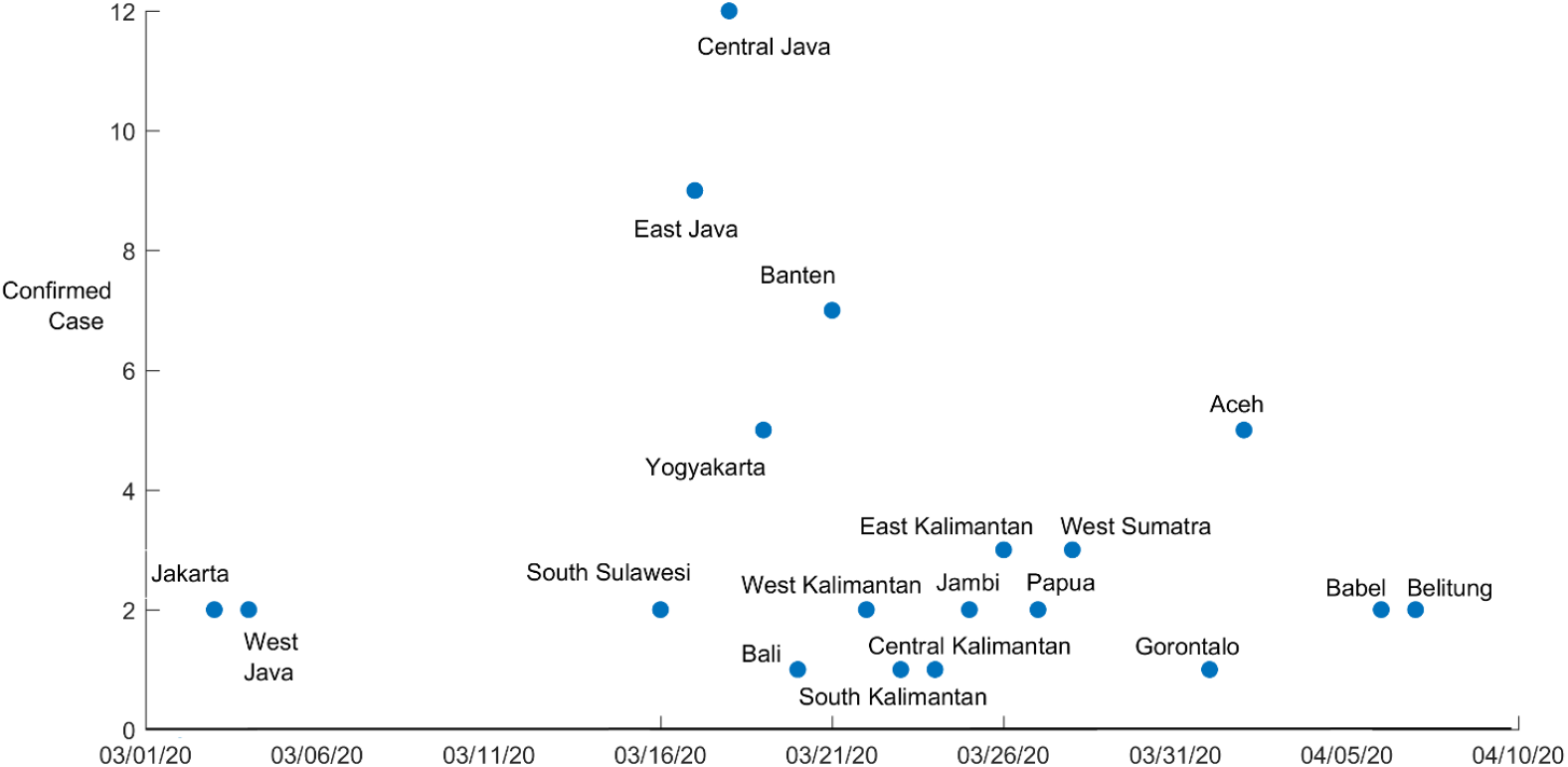
The beginning of the outbreak at many province in Indonesia

**Figure 4:**
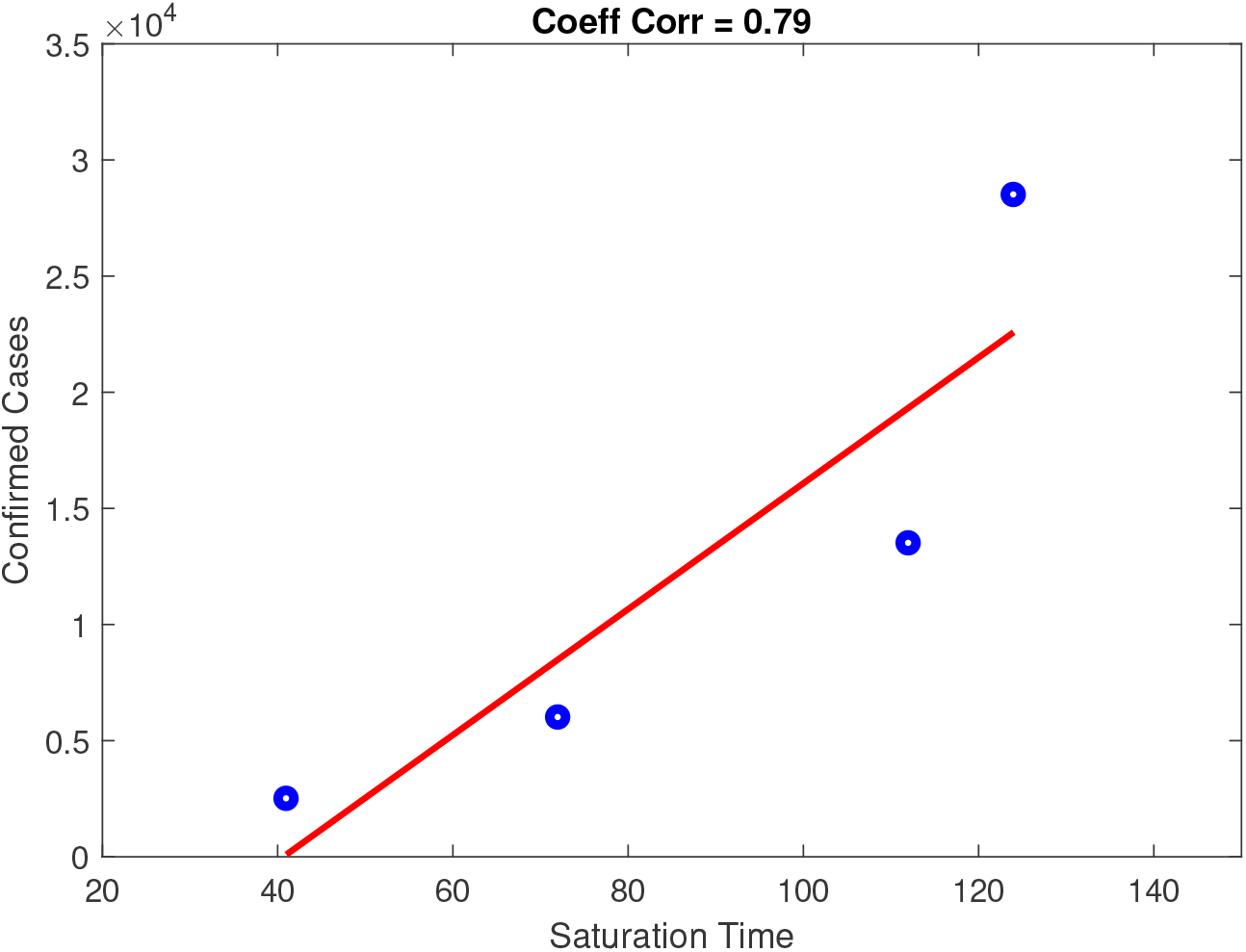
Correlation between time of saturation and confirmed cases

The coronavirus outbreak is finished when the confirmed cases reached the saturation. This condition may need a long time. Before conditions achieved, for economic reasons, the activity should be done by applying the protocol to prevent the transmission of the outbreak. We need the safety purpose put up with is called the condition of the new normal. To measure the security of an epidemic so defined basic reproduction number *r* = *β/γ* [6]. The value of *r <* 1 says that the potential for transmitting on other people has almost zero. This is called a safety condition. In our case, the basic reproduction number is depicted in Fig-5. The figure shows *r* decreases from 3 to 1 within 17 days. The result indicates that Indonesia is in a safe situation since 13 April 2020, but the fact is not. The use of *r* as an indication of safety is not right. Recent studies indicate that the bistable behavior may occur although the basic reproduction number is less than 1. This means that the policymakers do not make their decisions solely based on the guidance of the basic reproduction number only [14]. In this paper, we propose a new parameter to guidance the condition when the new normal begins. We introduce a parameter called receding defined as ℛ = *R* − *I*. This is depicted in Fig-6. At the time of the ℛ passes to the intersection between ℛ and *I*, we are in a situation where recovery person far more than infected people. In this case, we assume the situation can be controlled. On this condition, we can apply new normal but by taking into account the state of public health and health facilities. We should note that the ℛparameter is not the only way to look at the security but this is a guide when the basic reproduction number cannot be applied.

**Figure 5:**
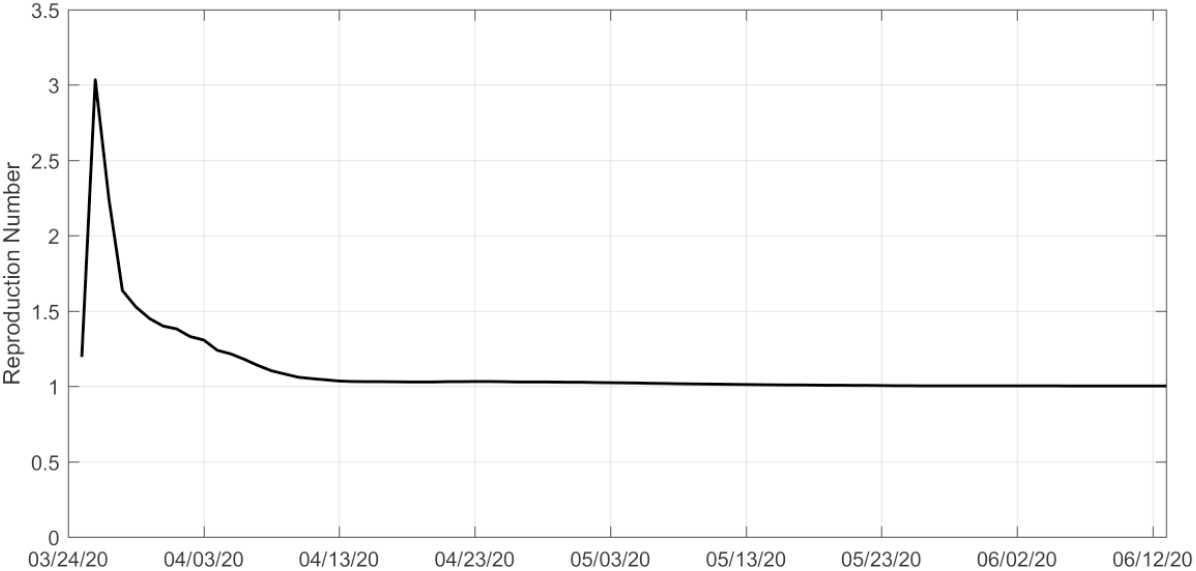
The time series of the basic reproduction number resulting from the calculation of the model from the beginning of the outbreak until June 14 2020

**Figure 6:**
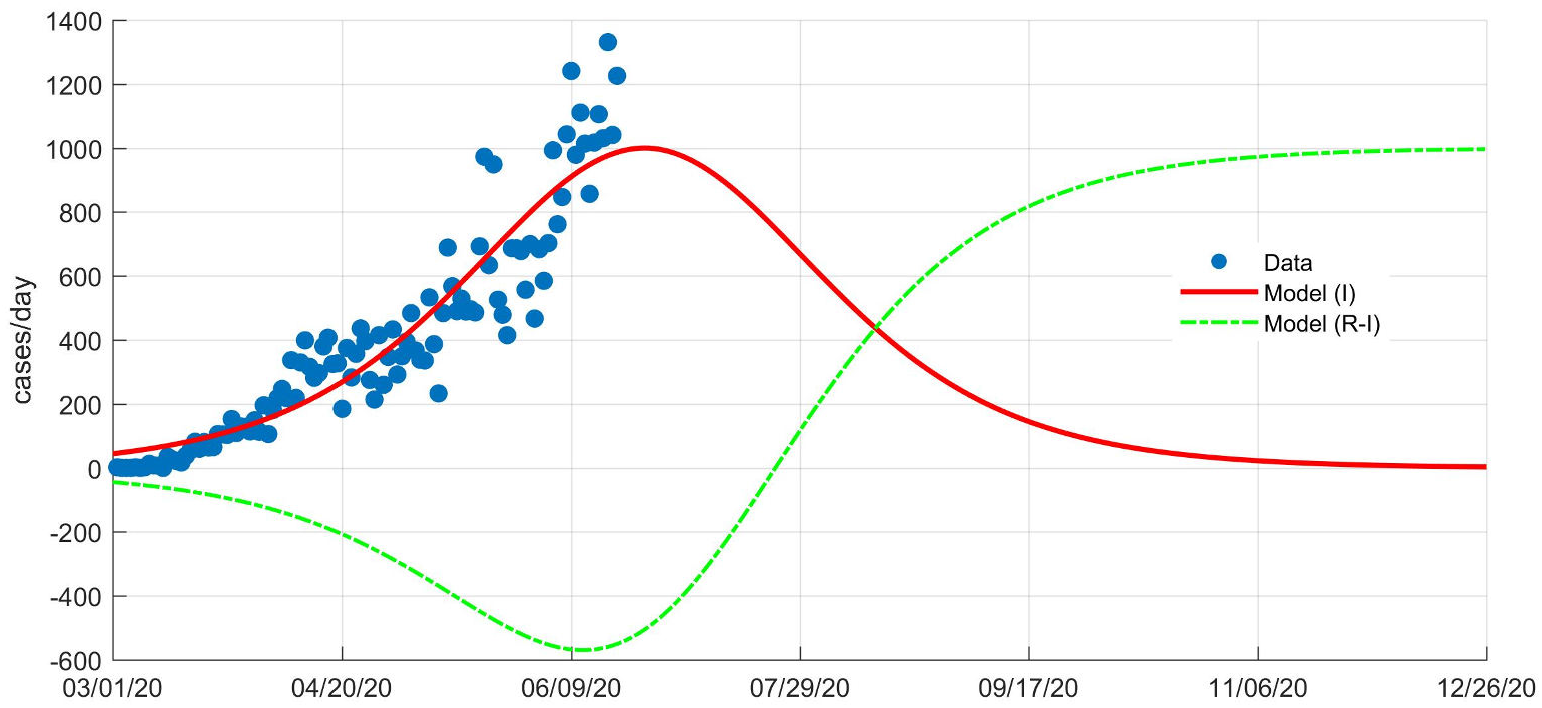
The receding parameter ℛ = *R* − *I* (dot-line), Infectious (red line) and case per day data (dot blue) from the calculation of the model from the beginning of the outbreak until June 14 2020. The intersection between ℛ and *I* line, stated a condition where a new normal can be implemented.

## 4 Summary

The dynamical analysis of the COVID-19 outbreak in Indonesia is investigated through the data-driven sir model with Matlab code developed by Batista. This model has shown that there are shifting the outbreak indicating the previous peak is specious. On the other hand, this model also explains the occurrence of a shifting of the time saturation gradually where it indicates that the acceleration of transmission is occurring in Indonesia. The data-driven SIR model can identify this serious condition that may be not known by another complex model. Introducing a receding parameter ℛ, we give a guidance when the new normal condition can be implemented. Due to Indonesia consisting of many islands, we got the data-driven SIR model for each island or province. In other words, we will deal with the n-compartments model where this is the focus research shortly.

## Data Availability

data can be obtained by Badan Nasional Penanggulangan Bencana Indonesia https://covid19.go.id/

## Acknowledgments

This research is funded by PTPSW-BPPT for fiscal years 2019/2020.

